# Involvement of fatigue in the effect of transcranial magnetic stimulation (TMS) on depression following COVID-19 and COVID-19 vaccination: a before-after study

**DOI:** 10.1101/2022.12.02.22282982

**Authors:** Ayane Kamamuta, Yuki Takagi, Mizuki Takahashi, Kana Kurihara, Hibiki Shibata, Kanata Tanaka, Katsuhiko Hata

## Abstract

**Background:** Patients recovering from COVID-19 often suffer long-term Long-COVID (e.g., depression, poor concentration, anxiety, sleep disturbances, and fatigue). Similar symptoms also rarely seem to occur after COVID-19 vaccination. There is still no effective treatment for these symptoms. We have had a clinical experience that patients presenting with psychiatric/physical symptoms due to COVID-19 or COVID-19 vaccination (defined as Long-COVID and Post-Vaccine patients) often recover after transcranial magnetic stimulation (TMS) and that TMS poorly heals depression in strongly fatigued patients.

**Aims:** 1. Determine whether there are differences in background characteristics and symptoms between Long-COVID and Post-Vaccine patients; 2. Examine whether TMS led to an improvement in their symptoms; 3. Test the involvement of fatigue in the recovery of depression of Long-COVID and Post-Vaccine patients with TMS.

**Methods:** We conducted a retrospective analysis using the medical records of the outpatient clinic of Tokyo TMS Clinic.

**Results:** 1. We found no differences in initial symptoms and courses of treatment between Long-COVID and Post-Vaccine patients. 2. All psychiatric/physical symptom scores after 10 TMS treatments were significantly better than before. Though these results are of before-and-after studies, numerous reports have suggested that TMS effectively improves depression, insomnia, anxiety, and related neuropsychiatric symptoms, which were also primary complaints of patients in this study. We thus attributed the improvement in QIDS, PHQ9 (Both indices of depression), and GAD7 (anxiety indicator) to TMS. 3. The recovery rate of depression in Long-Covid and Post-Vaccine patients with TMS decreased with the severity of fatigue.

**Conclusions:** This is the first report to elucidate the efficacy of TMS and the factors affecting it for psychiatric symptoms after COVID-19 and COVID-19 vaccination. Our study may lead to further validation of the effectiveness and mechanisms of TMS in patients suffering from Long-COVID and COVID-19 vaccine long-term adverse reactions.

## Indroduction

COVID-19 is raging and remains today’s most significant public health problem. As the number of people infected with COVID-19 has increased, so has the reporting of long-term symptoms (Long-COVID) in patients recovering from COVID-19. The symptoms of Long-COVID are diverse, with psychiatric and physical symptoms such as depression, poor concentration, anxiety, sleep disturbances, and fatigue making daily life difficult for patients after recovery^1-5^. Nevertheless, effective treatments for Long-COVID have not yet been identified, and doctors treating Long-COVID patients are exploring various treatment options.

Vaccination is essential to prevent the COVID-19 pandemic, which has been carried out on a large scale worldwide. Recently, long-term adverse reactions after COVID-19 vaccination with very low incidence have been reported ^6,7^. It includes many chronic conditions occurring even later than the neurological complications and thromboembolic/ thrombocytopenic events that sometimes happen within one month after the vaccination ^8,9^. It has been, therefore, difficult to be diagnosed ^7,10^. Only recently, the existence of COVID-19 vaccine long-term side effects is now being recognized, with an increasing number of reports suggesting that COVID-19 vaccine long-term adverse reactions may resemble the symptoms of Long-COVID ^11,12^. Yet, there are very few reports about the analysis of the symptoms of Long-COVID and COVID-19 vaccine long-term adverse reactions and effective treatments for them.

Transcranial magnetic stimulation (TMS) has been widely used in treating depression ^13-15^, bipolar disorder ^16^, obsessive-compulsive disorder ^17,18^, anxiety ^19,20^, insomnia ^21^, and neurological rehabilitation ^22,23^. So far, we have also performed TMS on patients with psychiatric symptoms such as depression, anxiety, insomnia, etc. Around the spring of 2020, the COVID-19 pandemic started to occur in Japan. Since January 2021, we have seen a gradual increase in outpatients complaining of psychiatric and physical symptoms lasting more than about a week, such as depression, poor concentration, anxiety, sleep disturbances, and fatigue after at least one week of SARS-CoV-2 infection (we define these patients as Long-COVID patients). Similarly, more outpatients since the summer of 2021 have complained of psychiatric/physical symptoms lasting more than about a week after at least one week of COVID-19 vaccination, such as depression, poor concentration, anxiety, sleep disorders, and fatigue. Although diagnosing that the COVID-19 vaccine causes these patients’ symptoms is difficult, it is true that the number of patients complaining that the COVID-19 vaccination causes their psychiatric/physical symptoms. So, we define them as Post-Vaccine patients. In our TMS treatments of Long-COVID and Post-Vaccine patients since January 2021, we have had clinical experiences that they often recover to the same degree as patients with typical depression, anxiety, and insomnia. We also had a clinical experience that patients appear to recover better from depression in Long-COVID and Post-Vaccine patients if they are less fatigued. As described in the Discussion section, an association between chronic fatigue and Long-COVID has also been suggested regarding pathogenic mechanisms ^24-26 27 28,29^.

To scrutinize these clinical empiricisms, we conducted a detailed retrospective analysis of the medical records of Long-COVID and Post-Vaccine patients to verify the following:

1. Are the symptoms of Long-COVID different from those of COVID-19 vaccine long-term adverse reactions?
2. The effectiveness of TMS for these patients’ clinical presentation.
3. Is fatigue involved in TMS-induced recovery from depression in Long-COVID and Post-Vaccine patients?

Again, it remains unclear if the symptoms of Long-COVID and COVID-19 vaccine long-term side effects are different and what effective treatments are available for them. We hope that the present study will lead to further validation of the efficacy of TMS for patients with symptoms of Long-COVID and COVID-19 vaccine long-term adverse reactions.

## Materials and Methods

### 1. Study design

This research is a retrospective study using medical records of patients who visited the Tokyo TMS Clinic. We compared four psychiatric/physical symptom test scores before and after the TMS treatment (before-and-after study). We analyzed the involvement of a chronic fatigue indicator at the initial visit in the recovery rate on the depression scale by Multivariate Covariance Analysis (MANCOVA).

### 2. Informed consent for patients

After explaining to patients the expected benefits and prospects of the treatment, costs, duration of treatment, anticipated side effects, and measures to deal with side effects, we obtained their consents in writing. We also brought approvals to provide the patients’ clinical data for academic research, including this study, to develop medicine, healthcare, and education.

### 3. Ethics committee approval

This study was conducted following the Declaration of Helsinki and approved by the BESLI CLINIC Ethics Committee (date of approval: 23 Oct 2022).

### 4. Transcranial magnetic stimulation device and coils

We performed transcranial magnetic stimulation with a Mag pro R30 (Magventure, Denmark) connected to a circular coil (Magventure cool-125).

### 5. TMS treatment

#### 5-1. Confirmation of indications for treatment

We asked the patient to complete a pre-interview form to ensure that the following absolute contraindications do not apply.

- Age below 18 years.
- Has a head injury.
- Hearing impairment.
- Pregnancy or possible pregnancy.
- Presence of metal (except titanium) near the stimulation site.
- Has a cochlear implant or implantable neurostimulator.
- Cardiac pacemaker.
- Spinal cord or ventricle with a spinal fluid shunt.
- Drug infusion device.
- Organic brain abnormalities found on MRI or CT.

A further examination by doctors confirmed that the patient was not eligible for TMS treatment, as listed below.

- History of psychiatric hospitalization.
- Strong feelings of hopelessness.
- Strong verbal abuse, violence, and irritability.
- Difficulty in communication.
- Recommendation for inpatient treatment.
- Schizophrenia
- Obsessive-compulsive disorder
- Personality disorders
- Somatoform disorders
- Flashbacks
- PTSD
- Attachment disorders
- Physical and mental conditions for which treatment at a specialized medical institution is recommended
- Seizure disorders such as epilepsy and severe physical illnesses. Patients who fall under the above checks or the doctor considers cannot be safely treated were excluded from treatment.

#### 5-2. The setting of the stimulation position

The physician selected the stimulation protocol at the first visit according to the patient’s symptoms. We asked the patients to sit comfortably and fit matching-sized treatment caps. We aligned the center of the hat with the mid-sagittal section and then measured the distance from the nasal root (nasion) to the median anterior margin of the lid. This measurement ensures reproducibility of the position of the head and cap after the second treatment by putting the cap on so that the distance between the anterior outer edge of the lid and the nasal root matches that of the first time. Next, the international 10/20 method, which is standardized in the positioning of electroencephalography (EEG) electrodes, was used to measure longitudinal (nasion to inion), transverse (tragus to tragus), and circumference (circumference). Based on these values, we calculated the distance along the circumference from the midline (X) and the distance from the vertex (Y) using the BeanF3 method. We thus determined the dorsolateral prefrontal cortex (DLPFC), which is the target of the treatment^30,31^.

#### 5-3. The setting of motor threshold (MT value)

The primary motor cortex of the abductor pollicis brevis muscle (APB) ipsilateral to the stimulation position was used as the reference. We defined the MT value as the minimum stimulus intensity at which the APB muscle contraction could be visually confirmed at least five times out of 10 by single-shot stimulation (5 Hz).

#### 5-4. Stimulus intensity

After setting the stimulation position and motor threshold, the dorsolateral prefrontal cortex (DLPFC), which is the target of the treatment, is determined by gradually increasing the output from 50% of the motor threshold while confirming the patient’s pain (e.g., when MT is 50, stimulation intensity of 40 is 80% of MT). If the patient felt intense pain, we set the initial stimulation intensity at 60-80% of the MT value. We increased the output as the number of treatments increased to 120% of the MT value indicated in the Guidelines for the Appropriate Use of Repetitive Transcranial Magnetic Stimulation (The Japanese Society of Psychiatry and Neurology).

#### 5-5. Start of treatment

We asked the patient to sit in a comfortable position and aligned the center of the cap used for measurement at the initial visit with the mid-sagittal section. We then measured the distance from the nasal root (nasion) to the median anterior margin of the cap and matched it to the cap position at the initial visit. The patient was asked to wear earplugs to reduce the stimulation sound. We set the coil at the stimulus position and set the treatment parameters.

#### 5-6. Assessment of safety and treatment efficacy

QIDS (Quick Inventory of Depressive Symptomatology) ^32^, PHQ9 (Patient Health Questionnaire-9) ^33^, GAD7 (Generalised anxiety disorder-7) ^34^, PS (Performance Status) ^35,36^ and other standardized scales were used to determine treatment efficacy, side effects, and future treatment strategy.

### 6. Data collection

We selected medical records of patients with post-COVID-19 sequelae and long-term COVID-19 vaccine side effects as the main complaint from approximately 2000 outpatients between 15 January 2021 and 29 September 2022 (Long-covid group: 100 cases, Post-Vaccine group: 29 cases, 129 cases in total: Figure 1). We excluded patients who were judged to be off-label for treatment regarding the Guidelines for the appropriate use of rTMS (Japanese Society of psychiatry and neurology) or who refused to accept treatment after the explanation of the TMS procedure (Long-covid group: 11 patients, Post-Vaccine group: 5 patients, a total of 16 patients). We excluded patients who received TMS only for the first time because they only wanted to try the TMS procedure (Long-covid group: 11 patients, Post-Vaccine group: 5 patients, 27 patients in total). We excluded patients who discontinued treatment before the end of 10 TMS sessions (between the 2nd and 10th sessions: Long-covid group: 21 patients, Post-Vaccine group: 5 patients, 26 patients in total). Consequently, we included in the analysis 46 cases with post-COVID-19 sequelae as the main complaint and 14 patients with long-term COVID-19 vaccine side effects as the main complaint with medical records at the initial visit and after 10 TMS treatments.

**Figure 1.**
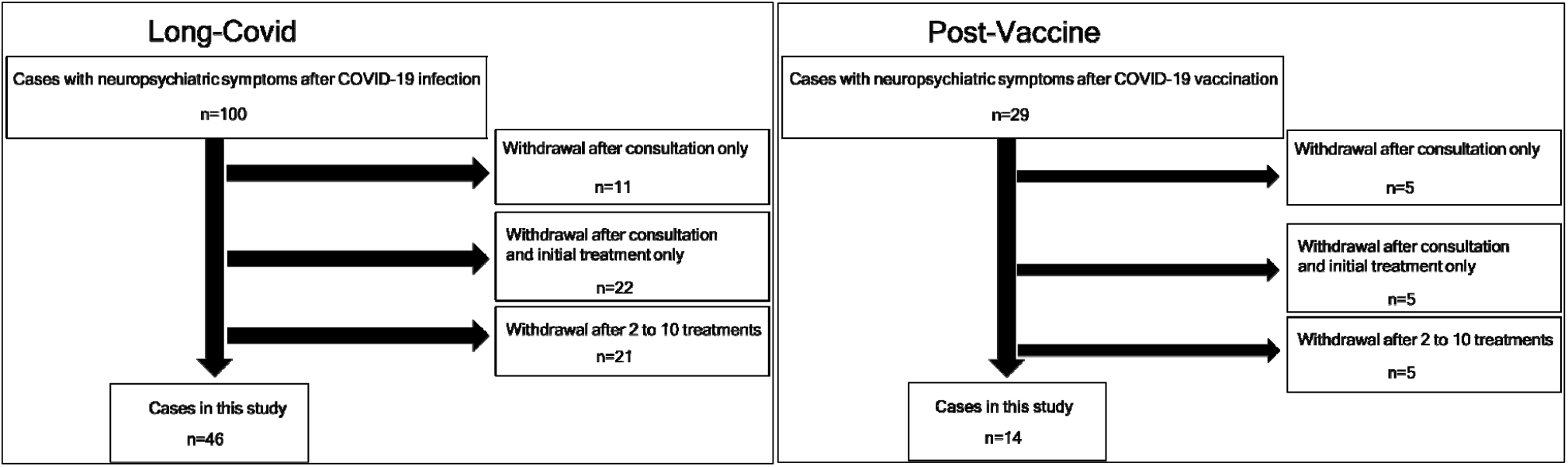
List of Target Data and Exclusions

We defined the Long-Covid group as patients who, at the first visit, claimed to have psychiatric/physical symptoms applicable to QIDS, PHQ9, GAD7, and PS lasting approximately one week or more after at least one week of COVID-19 and who thought the disease caused them. We defined the Post-Vaccine group as patients who claimed psychiatric/physical symptoms applicable to QIDS, PHQ9, GAD7, and PS lasting approximately one week or more after at least one week of COVID-19 vaccination and who thought the disease caused them. Therefore, the present study did not conduct rigorous causal scrutiny of whether SARS-COV-2 infection or its vaccination elicited their symptoms.

Items extracted from the medical records include chief complaint, sex, magnetic stimulation intensity (% of motor threshold (MT)), rTMS protocol, medication use, duration from the first visit to 10 TMS treatments, psychiatric/physical test scores (QIDS, PHQ9, GAD7, PS) at the initial visit and after of tenth TMS treatments. Here, the chief complaint is the basis for determining whether the patient falls into the Long-COVID or Post-Vaccine group. The rTMS protocol includes high-frequency rTMS (left DLPFC), low-frequency rTMS (right DLPFC), and other protocols.

### 7. Outcomes

The primary outcome was the partial regression coefficient of PS at the first visit in MANCOVA with the improvement rate of QIDS with TMS (ΔQIDS) as the dependent variable. Secondary outcomes were QIDS, PHQ9, GAD7, and PS at the initial visit and after 10 TMS procedures and their improvement rates.

### 8. statistical analysis

Among items extracted from the medical records, we defined patient background factors as follows: sex, age, TMS intensity, TMS protocol, presence or absence of medication, and the number of days taken from the first visit to 10 treatments. The Wilcoxon test verified the difference in each background factor between the Long-COVID and Post-Vaccine groups. QIDS scores ranged from 0 to 27, PHQ9 from 0 to 27, GAD7 from 0 to 21, and PS from 0 to 9, with higher scores indicating more severe symptoms. We calculated QIDS (%), PHQ9 (%), GAD7 (%), and PS (%) using the following formulas.

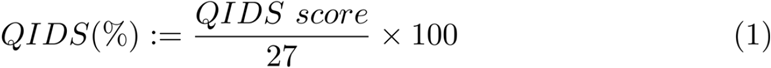

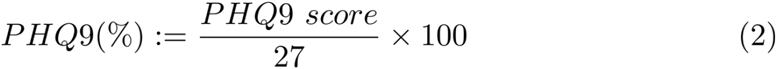

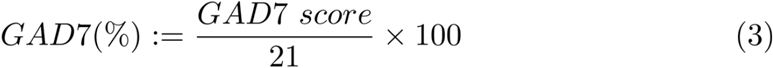

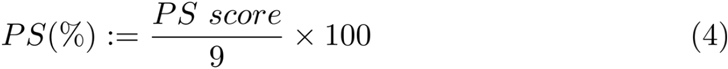

We calculated ΔQIDS, ΔPHQ9, ΔGAD7, and ΔPS as follows.

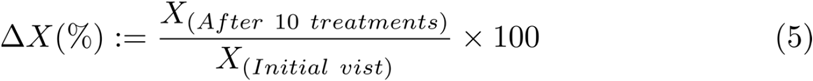

Note that X is substituted for QIDS, PHQ9, GAD7, and PS. We excluded data with a psychiatric/physical symptom test score of zero at the first visit when calculating ΔX. We performed univariate analyses in Tables 1 and 4 and used the Wilcoxon test for significance analyses. We performed univariate analyses in Tables 2 and 3 and used the Wilcoxon signed rank test for significance checks. When examining the multivariate correlations for each psychiatric/physical symptom test score (Figures 2 and 3), we determined the correlation coefficient, the p-value of the correlation, and the scatterplot matrix. We set the number of significant digits for each data to two decimal places and the p-value to one significant digit. REML estimation was used to handle missing data in the multivariate correlation analysis.

**Table 1.** Characteristics of the patients

|  |  | Long-Covid (N=46) | Post-Vaccine (N=14) | P value |
| --- | --- | --- | --- | --- |
| Male(%) |  | 31(67.39) | 6(42.86) | - |
| Age(years old) |  | 39.5(29-47.25) | 34(25.25-42) | - |
| Stimulus intensity of TMS (% of MT) |  | 91(80-100) | 84.5(69.75-97.75) | - |
| Stimulation Protocol | High frequency rTMS (left DLPFC)(%) | 41(89.13) | 13(92.86) | - |
|  | Low frequency rTMS (right DLPFC)(%) | 4(8.70) | 0(0.00) | - |
|  | Other(%) | 1(2.17) | 1(7.14) | - |
| Medication(%) |  | 28(60.87) | 12(85.71) | - |
| Duration of treatment(days) |  | 12(7.75-19.5) | 5.5(3.75-22.75) | - |

Data are presented as median (IQR) for continuous measures and n (%) for categorical measures.

**Table 2.** Differences in each psychiatric/physical symptom test score at the first visit.

|  |  |  |  |  |  |  |
| --- | --- | --- | --- | --- | --- | --- |
| Long-Covid |  |  |  |  |  |  |
|  | PHQ9(%)<br>QIDS(%) | GAD7(%)<br>QIDS(%) | GAD7(%)<br>PHQ9(%) | PS(%)<br>QIDS(%) | PS(%)<br>PHQ9(%) | PS(%)<br>GAD7(%) |
| N | 46 | 46 | 46 | 39 | 39 | 39 |
| Difference(%) | 18.52(7.41,26.85) | 5.29(-1.98,17.59) | -11.11(-24.07,2.91) | 3.70(-7.41,25.93) | -14.81(-22.22,3.70) | 3.17(-9.52,20.63) |
| P value | <.0001* | 0.005* | <.0001* | 0.04* | 0.02* | 0.5 |
| Post-Vaccine |  |  |  |  |  |  |
|  | PHQ9(%)<br>QIDS(%) | GAD7(%)<br>QIDS(%) | GAD7(%)<br>PHQ9(%) | PS(%)<br>QIDS(%) | PS(%)<br>PHQ9(%) | PS(%)<br>GAD7(%) |
| N | 14 | 14 | 14 | 12 | 12 | 12 |
| Difference(%) | 22.22(2.78,34.25) | 2.91(-6.61,23.94) | -8.99(-23.81,3.57) | 25.93(8.33,50.93) | 11.11(-9.26,28.70) | 11.90(-3.57,49.60) |
| P value | 0.002* | 0.2 | <0.05* | 0.005* | 0.3 | 0.06 |

Data are presented as median (IQR). \*Significant (P<0.05)

**Table 3.** Changes in each psychiatric/physical symptom test score between the first visit and after 10 TMS treatments

| Psychiatric Symptom |  | Number | Scores | P value |
| --- | --- | --- | --- | --- |
| QIDS (range 0-27) | Initial visit | 46 | 6(3-8) | <.0001* |
|  | After 10 treatments | 46 | 3(1-5) |  |
| PHQ9(range 0-27) | Initial visit | 46 | 10(7-14) | <.0001* |
|  | After 10 treatments | 46 | 5(3-9) |  |
| GAD7(range 0-21) | Initial visit | 46 | 5(3-10) | <.0001* |
|  | After 10 treatments | 46 | 3(1-6) |  |
| Physical Symptom |  |  |  |  |
| PS(range 0-9) | Initial visit | 39 | 2(1-4) | 0.01* |
|  | After 10 treatments | 39 | 1(1-3) |  |

Post-Vaccine
| Psychiatric Symptom |  | Number | Scores | P value |
| --- | --- | --- | --- | --- |
| QIDS (range 0-27) | Initial visit | 14 | 7(2.75-9) | 0.04* |
|  | After 10 treatments | 14 | 5.5(1-7.5) |  |
| PHQ9(range 0-27) | Initial visit | 14 | 14(3.75-17.25) | 0.0002* |
|  | After 10 treatments | 14 | 9(2.75-11.75) |  |
| GAD7(range 0-21) | Initial visit | 14 | 5.5(1-12.25) | 0.001* |
|  | After 10 treatments | 14 | 3.5(0-7.25) |  |
| Physical Symptom |  |  |  |  |
| PS(range 0-9) | Initial visit | 12 | 5.5(2-6.75) | 0.02* |
|  | After 10 treatments | 12 | 4.5(1.25-5.75) |  |

Scores are presented as median (IQR). \*Significant (P<0.05)

**Table 4.**
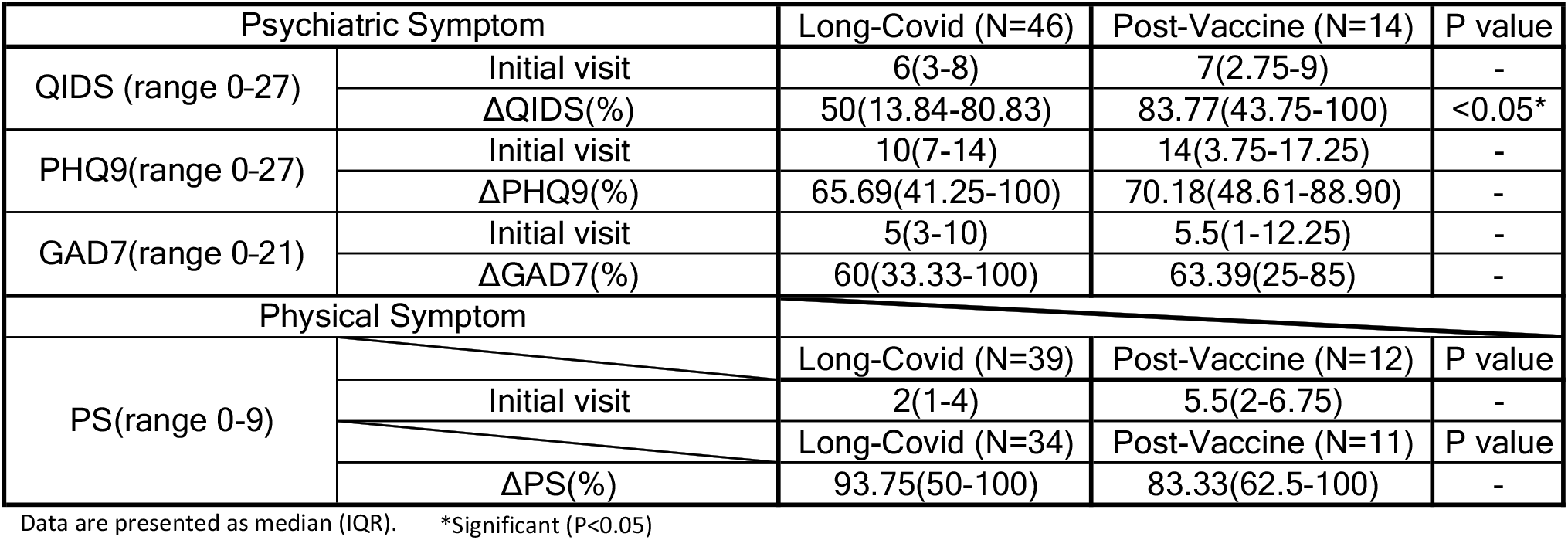
Comparison of Long-Covid and Post-Vaccine patients on the initial value of each psychiatric/physical symptom test and the improvement rate with TMS treatment.

**Figure 2.**
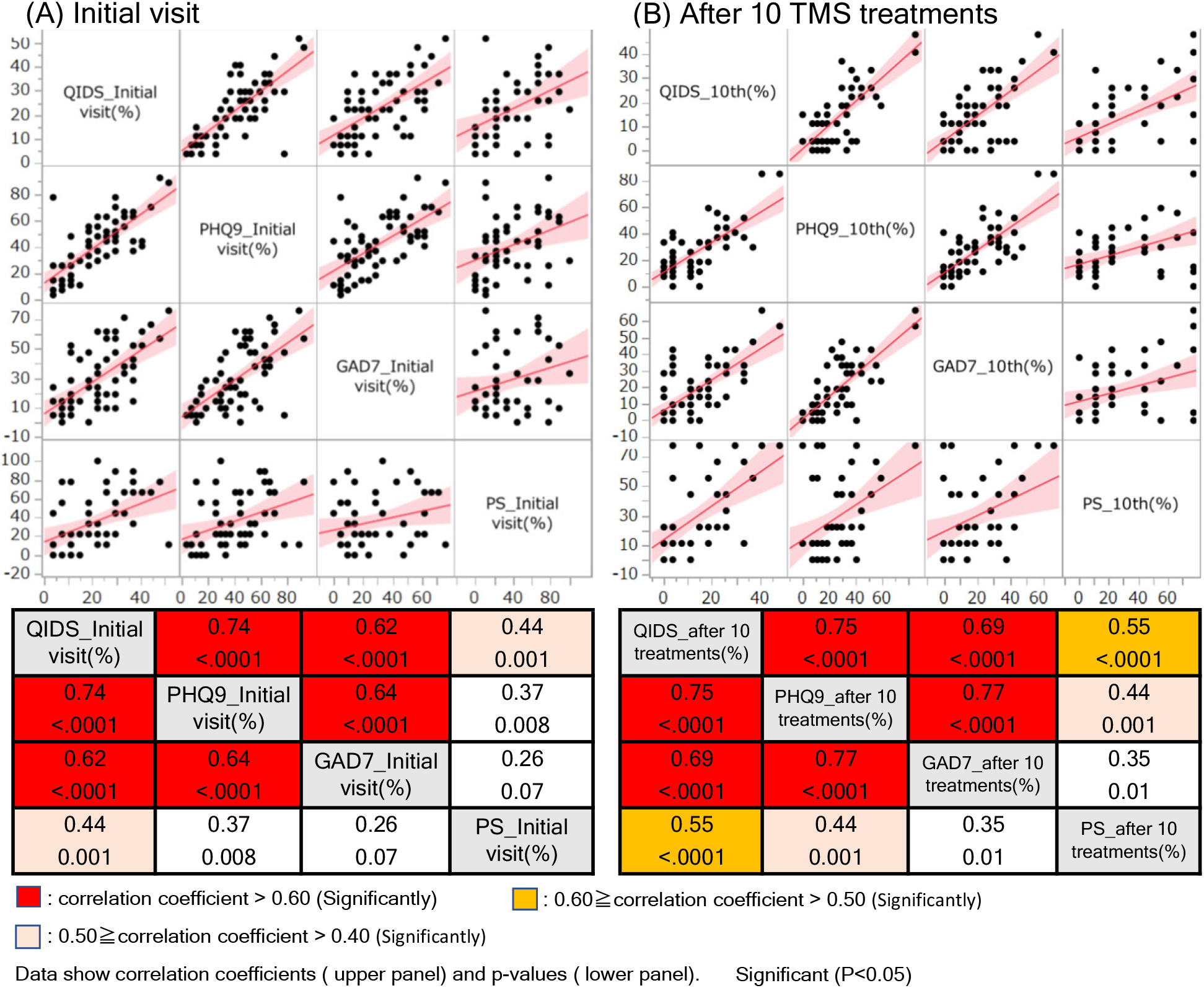
Correlations between each psychiatric/physical symptom test score.

**Figure 3.**
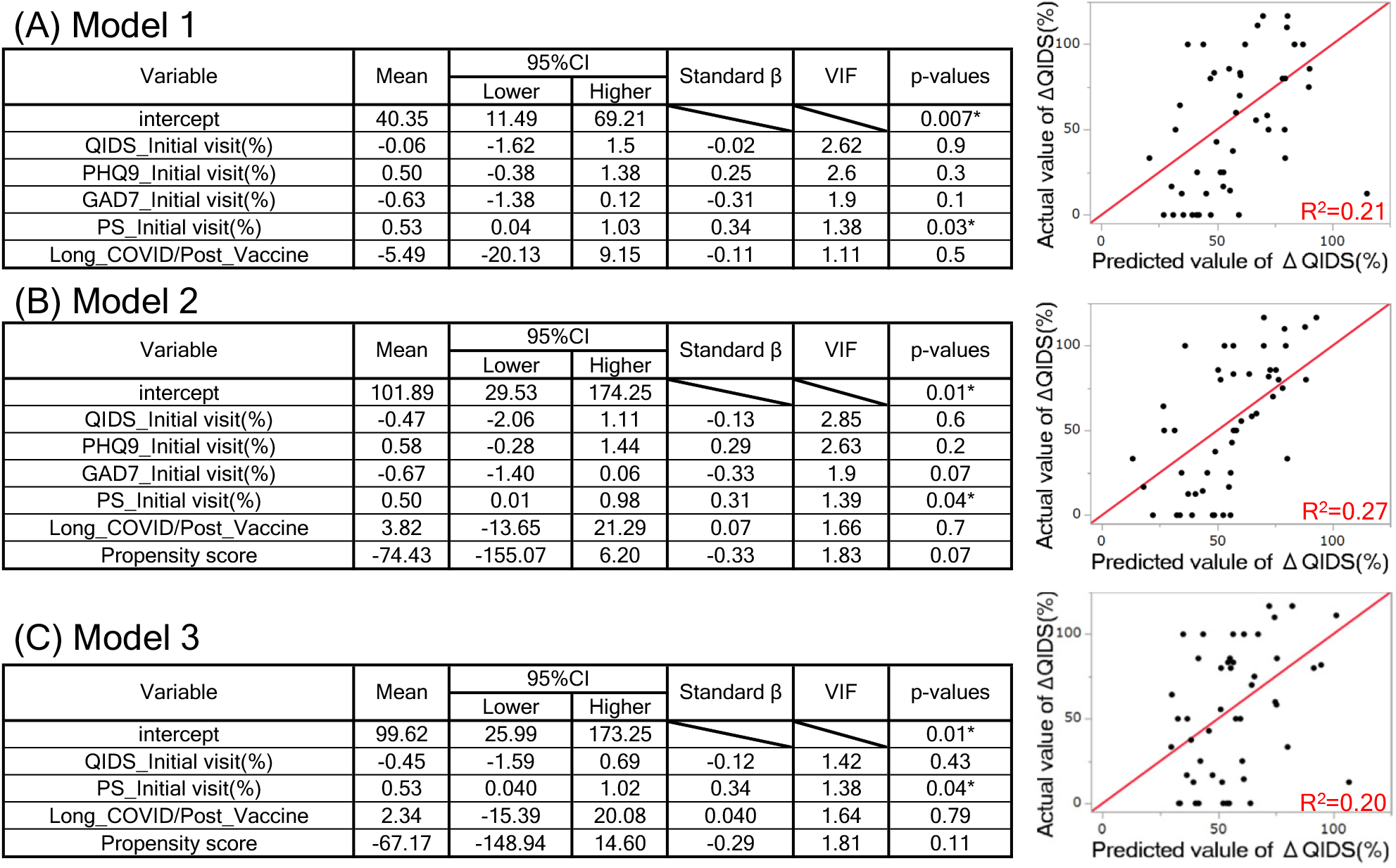
Multivariate analysis of covariance between ΔQIDS (%) and each psychiatric/physical symptom test score at first visit.

We conducted single regression analyses and a MANCOVA to identify initial visit scores correlated with the primary outcome, ΔQIDS, and the secondary outcome, the percentage improvement in PHQ9, GAD7, and PS. When carrying out the MANCOVA with ΔQIDS as the dependent variable, we used as independent variables the four initial QIDS, PHQ9, GAD7, PS, and information on whether the patient belonged to the Long-Covid or Post-Vaccine group, which was made into a dummy variable, with Long-Covid=1, Post-Vaccine=-1.

For undertaking a MANCOVA using propensity scores, we obtained a propensity score by logistic regression analysis with the independent variables of age, gender, medication status, TMS intensity, TMS protocol, and duration of treatment, and the dependent variable of Long-Covid/Post-Vaccine. We then performed a MANCOVA on the independent variables of this propensity score plus initial visit QIDS, PHQ9, GAD7, PS, and dummy variable of Long-Covid/Post-Vaccine, as the dependent variable of ΔQIDS (%). To avoid overfitting, we set the maximum number of independent variables for MANCOVA to six ^37^.

We performed all statistics using JMP pro-version 15.0.0 (SAS Institute Inc., Cary, NC, USA) and considered p <0.05 (two-tailed) statistically significant.

## Results

Once again, let us review the definitions of the terms used in this study. We defined “long-COVID patients” as outpatients who have complained of psychiatric/physical symptoms lasting more than about a week, such as depression, poor concentration, anxiety, sleep disturbances, and fatigue after at least one week of COVID-19. Similarly, outpatients who have complained of psychiatric/physical symptoms lasting more than about a week after at least one week of COVID-19 vaccination, such as depression, poor concentration, anxiety, sleep disorders, and fatigue, were defined as “Post-Vaccine patients”. Therefore, we did not conduct rigorous causal scrutiny of whether these symptoms were evoked by SARS-COV-2 infection or its vaccination. The psychiatric/physical symptom tests used in this study were QIDS, PHQ9, GAD7, and PS. The QIDS and PHQ9 are indicators of depression ^32,33^ and the GAD7 of anxiety disorder ^34^. We used PS as an indicator of fatigue symptoms. PS is one of the reference indicators used in the criteria created by the Japanese Ministry of Health, Labour and Welfare (MHLW) based on the Myalgic Encephalomyelitis/Chronic Fatigue Syndrome (ME/CFS) diagnostic criteria, which were developed based on the requirements set by the Institute of Medicine (IOM), now the National Academy of Medicine (NAM) in the USA, in 2015.

First, we examined the background of Long-Covid and Post-Vaccine patients to determine which characteristics patients presented to the clinic and if there were any significant differences between the two groups. Table 1 shows no significant differences between the two groups regarding gender, age, TMS stimulation intensity, protocol, medication, and the number of days taken from the first visit to the 10th treatment.

We examined which items tended to be higher in each psychiatric/physical symptom test at the first visit in Long-Covid and Post-Vaccine patients (Table 2). As the highest score for each test differed, we normalized each test according to equations (1) to (4). We then examined for significant differences between each psychiatric/physical symptom test. We found that both Long-Covid and Post-Vaccine patients showed the same trend (Table 2). That is, both Long-Covid and Post-Vaccine patients showed significantly higher values for PHQ9>GAD7, PS>QIDS, in that order.

Using the Wilcoxon signed rank test, we examined how each psychiatric/physical symptom test score changed before and after ten sessions of TMS treatment. The results showed that QIDS, PHQ9, GAD7, and PS improved significantly in both the Long-Covid and Post-Vaccine groups (Table 3). We should note here that this is a retrospective before-and-after comparison study. However, numerous reports have shown that TMS effectively improves depression, insomnia, anxiety, and their associated neuropsychiatric symptoms ^13-15,19-21^. We have also experienced an improvement in these patients with TMS. In the present study, Long-COVID and Post-Vaccine patients had varying degrees of these symptoms as their primary complaints. Therefore, it is likely inferred that QIDS, PHQ9, and GAD7 have improved with TMS. However, the level of evidence is low concerning TMS of chronic fatigue, although positive results have been reported ^38^. So, we cannot conclude from these results alone that the improvement in PS is due to TMS.

We conducted a univariate analysis of the initial psychiatric/physical symptom test score and the improvement rate for each score between Long-Covid and Post-Vaccine patients. To calculate the improvement rate, we divided the score after 10 TMS procedures by the score at the first visit, as shown in equation (5) ΔX (%), where lower values indicate a better improvement rate. As shown in the “Initial Visit” row of each psychiatric/physical symptom test in Table 4, there were no significant differences in the scores between Long-Covid and Post-Vaccine patients at the initial visit. This result is similar to the conclusions drawn from Table 2. Regarding the rate of improvement, only ΔQIDS differed between the two groups (row “ΔQIDS (%)” in Table 4). At first glance, Long-Covid patients seem to have a significantly better improvement rate in QIDS than Post-Vaccine patients. However, the Multivariate Covariance Analysis (MANCOVA) described below (Figures 3A, B, and C) no longer detected this difference. We thus found no significant differences in symptoms and improvement rates between Long-Covid and Post-Vaccine patients at the first visit (Tables 2 and 4). These results agree with our clinical empiricism that symptoms of Long-Covid and Post-Vaccine patients are very similar. Therefore, we assume that the initial signs and their course are the same in Long-Covid and Post-Vaccine patients.

We investigated the degree of correlation between the psychiatric/physical test scores at the initial visit and after TMS (Figures 2A and B). Not surprisingly, the correlations between psychological scores (QIDS, PHQ9, and GAD7) were high both at the initial visit and after TMS. Interestingly, QIDS vs. PS was correlated both at the first visit and after TMS, and PHQ9 vs. PS after TMS. These results suggest that, in line with our hypothesis, fatigue may affect the recovery rate from Long-Covid and Post-Vaccine depression (hereafter referred to as “Covid-related depression”).

We explored the impact of initial fatigue symptoms on the improvement rate of Covid-related depression using single regression analysis and MANCOVA. In both studies, we found that ΔQIDS (%), the rate of improvement in QIDS, was significantly positively correlated with PS (%) at the initial consultation (Figure 3A). No significant correlations were obtained except for this combination, by the way. We conducted a MANCOVA with as dependent variable ΔQIDS (%), as independent variables each of the four initial visiting psychiatric/physical test scores and a dummy variable for the information about the group the patient belongs to (Long-Covid/Post-Vaccine) (Figure 3A: Model 1). Then, PS (%) at the first visit showed a significant positive partial regression coefficient (Figure 3A). To further investigate the influence of PS (%) on ΔQIDS (%) at the initial visit, we carried out a MANCOVA with as many confounders for Long-Covid/Post-Vaccine as possible recast as a single variable (propensity score). We obtained propensity scores by logistic regression analysis with age, gender, medication status, magnetic stimulation intensity, magnetic stimulation protocol, and duration of treatment (shown in Table 1) as independent variables and Long-Covid/Post-Vaccine as a dependent variable (Figure 3B). We conducted a MANCOVA with as independent variables this propensity score as well as Long-Covid/Post-Vaccine, QIDS at the first visit, PHQ9, GAD7, and PS, as a dependent variable ΔQIDS (%) (Figure 3B: Model 2). Note that we considered it reasonable to include six independent variables in the present MANCOVA concerning previous studies ^37^. Similar to Figure 3A, PS (%) at the first visit showed a significantly positive partial regression coefficient, even considering many confounds (Figure 3B). Finally, we adjusted for multicollinearity between the independent variables and performed a MANCOVA. Models 1 and 2 show the variance inflation factor (VIF) between the QIDS, PHQ9, and GAD7 at the first visit was around 2 to 3, as high correlation coefficients between these psychological test scores are also indicated in Figure 2. We thus conducted a MANCOVA of ΔQIDS (%) by selecting only QIDS from the psychological test scores as the independent variable and adding PS at initial diagnosis, Long-Covid/Post-Vaccine, and propensity score from Model 2 as independent variables (Figure 3C: Model 3). The results also showed a significant positive partial regression coefficient for PS (%) at the initial consultation.

As mentioned in the results of Table 4, Long-Covid/Post-Vaccine did not significantly affect ΔQIDS (%). These results suggest that Covid-related depression is less likely to improve with TMS in patients with more pronounced fatigue symptoms.

## Discussion

The present study is the first report to elucidate the validity of TMS and the factors underlying it for psychiatric symptoms after COVID-19 and its vaccination. We found no difference in characteristics or initial symptoms between patients who presented with psychiatric/physical symptoms after COVID-19 (Long-Covid patients) and those who presented with psychiatric/physical symptoms after COVID-19 vaccination (Post-Vaccine patients). Comparing QIDS, PHQ9, GAD7, and PS before and after TMS treatment in both groups, all items were significantly improved. There were no significant differences between both groups in the rate of improvement of QIDS, PHQ9, GAD7, and PS with TMS. We, therefore, assumed that the initial symptoms and course of Long-Covid and Post-Vaccine patients were the same and defined their depression as “Covid-related depression”. Also, though this is a retrospective, before-and-after study, numerous reports suggest that TMS may have improved QIDS, PHQ9, and GAD7. We thus attributed the improvement in QIDS, PHQ9, and GAD7 to TMS. Meanwhile, these results alone made it impossible to conclude that TMS improved PS. We then explored fatigue influences on the improvement rate of Covid-related depression by single regression analysis and MANCOVA. We found that the higher the PS at the initial visit, i.e., the stronger the chronic fatigue symptoms, the worse the Covid-related depression recovery rate.

Long-Covid and Post-Vaccine patients had significantly higher psychiatric/physical symptom values at their first visit in the order PHQ9>GAD7, PS>QIDS, and they showed similar trends (Table 2). In fact, depression, poor concentration, sleep disturbances, anxiety, and fatigue were common to both groups of patients visiting the Tokyo TMS Clinic. The recovery rate of each symptom also did not differ between the two groups (Table 4). COVID-19 is reported to cause severe inflammatory symptoms, and even minor infections are associated with cytokine elevation and brain microglial activation that persists for a long time ^27^. On the other hand, the COVID-19 vaccine, including mRNA vaccines distributed in Japan (mRNA-1273 SARS-CoV-2 Vaccine and BNT162b2 mRNA COVID-19 Vaccine ^39,40^), is known to cause Long-Covid-like symptoms, albeit at a low rate^11^. A recent report suggested that the COVID-19 vaccine causes severe inflammatory symptoms by disrupting innate immunity by suppressing IFN-α signaling, failing the system to prevent and detect intracellular malignant transformation, and generating large numbers of exosomes carrying spiked glycoproteins ^12^. Thus, the similarity in pathogenesis mechanisms between the Long-Covid and Post-Vaccine patients may have led to similar symptoms in both groups.

Even though both the QIDS and PHQ9 are highly sensitive and specific diagnostic criteria for depression, both Long-Covid and Post-Vaccine patients tended to score significantly higher on the PHQ9 than on the QIDS at the first visit ^32,33^. It is necessary to conduct further research on whether this difference depends on how the QIDS and PHQ9 questions are asked or on the symptoms of the Long-Covid/Post-Vaccine patients.

We found that after 10 TMS treatments for patients presenting with psychiatric/physical symptoms, including depression, triggered by COVID-19 and COVID-19 vaccination, there was a significant improvement in all test scores compared to pre-treatment (Table 3). It is impossible to conclude from this before-and-after study alone that TMS ameliorates psychiatric/physical symptoms. However, several reports have demonstrated the therapeutic effects of TMS on depression, insomnia, anxiety, and their associated neuropsychiatric symptoms ^13-15,19-21^. The patients in this study who claim COVID-19 or COVID-19 vaccination as the cause also have chief complaints of psychiatric symptoms, including depression, poor concentration, insomnia, and anxiety. The depression scales QIDS and PHQ9 used in this study include questions on depression, poor concentration, and insomnia. The GAD7 is a diagnostic indicator of the degree of generalized anxiety disorder. It would therefore be natural to consider that TMS improved QIDS, PHQ9, and GAD7 ^32-34^. On the other hand, there is still little medical evidence on the effectiveness of TMS for chronic fatigue ^38^. In this study, although TMS significantly improved PS compared to pre-treatment, we cannot rule the possibility out that PS could have improved spontaneously even without TMS. Whether TMS improves fatigue symptoms remains to be further explored by conducting a double-blind and randomized controlled trial.

We found by MANCOVA that the higher the PS (used as a reference measure for ME/CFS diagnosis) at the initial diagnosis, the worse the improvement rate of QIDS. Although it is impossible to conclude that a person has ME/CFS solely because of a high PS, the results are consistent with our clinical experience, suggesting that chronic fatigue may influence the improvement of Covid-related depression. The association between chronic fatigue and Covid-related depression is also indicated in terms of both pathogenic mechanisms. Patients with ME/CFS have higher levels of oxidative stress and widespread microglial activation in the brain compared to healthy subjects ^24,25^. Interestingly, the level of microglial activation in ME/CFS patients positively correlated with cognitive impairment and the severity of depression and pain. On the other hand, autopsy reports of patients who died from COVID-19 reported activation of microglia in the brain in cases in which SARS-COV-2 infection in the brain was not confirmed ^26^. Experimental studies in mice with SARS-COV-2 infection confined to the lungs also found microglia activation in the brain’s white matter ^27^. A recent study found that Long-Covid patients are subjected to higher oxidative stress than healthy subjects ^28^. These findings suggest that the activation of microglia in the brain and an increase in oxidative stress evoke ME/CFS and Long-Covid. Brain inflammation and high oxidative stress in patients with high PS levels could be linked to resistance to treatment for TMS. Controlling inflammation and oxidative stress in such patients appears necessary for recovery from Covid-related depression with TMS ^29^.

Limitations of this study include the following. Improvements with TMS in psychiatric/physical symptoms were obtained by a before-and-after comparative study. We could not test which TMS stimulation protocols produced recovery of Covid-related depression, as the physician selected the stimulation protocols at the first visit according to the patient’s symptoms. Most of the stimulation protocols were high-frequency stimulation of the left DLPFC. We cannot rule out the possibility that uncorrected and unknown confounding factors may have influenced the results when performing MANCOVA.

## Data Availability

All data generated in this study are available if the BESLI CLINIC Ethics Committee considers it a reasonable request.

## Author contributions

K.H. conceived the ideas and compiled this study. A.K., K.K., H.S., and K.T. operated the TMS equipment. A.K., Y.T., M.T., K.K., H.S., K.T., and K.H. collected medical records. A.K. extracted data for analysis from medical records and summarized the TMS practice. K.H. analyzed the data statistically. A.K., Y.T., K.T., and K.H. discussed the statistical analysis results from a medical perspective. K.H. prepared the manuscript. All authors reviewed the manuscript.

## Acknowledgments

We thank BESLI Medical Corporation, all the staff at the Tokyo TMS Clinic, and all the patients who cooperated in the study. We gratefully acknowledge the support of the JSPS Grants-in-Aid for Scientific Research No. JP18K10858 and 21H03327 and the Watanabe Foundation for supporting this research.

